# Impact assessment of mobility restrictions, testing, and vaccination on the COVID-19 pandemic in India

**DOI:** 10.1101/2022.03.24.22272864

**Authors:** Jeonghyun Shin, Quynh Long Khuong, Kaja Abbas, Juhwan Oh

## Abstract

**Background:** Before the availability of vaccines, countries largely relied on mobility restriction and testing to mitigate the COVID-19 pandemic. Our aim is to assess the combined impact of mobility restriction, testing, and vaccination on the COVID-19 pandemic in India.

**Methods:** We conducted a multiple regression analysis to assess the impact of mobility, testing, and vaccination on COVID-19 incidence between April 28, 2021 to November 24, 2021 using data from Our World in Data and Google Mobility Report. The 7-day moving average was applied to offset the daily fluctuations in the mobility and testing. Each independent variable was lagged to construct a temporal relationship, and waning vaccination efficacy was taken into consideration. We performed additional analysis for three time periods between March 28, 2020 to November 24, 2021 (1^st^: March 28, 2020 ∼ October 7, 2020, 2^nd^: October 8, 2020 ∼ April 27, 2021, 3^rd^: April 28, 2021 ∼ November 24, 2021) to examine potential heterogeneity over time.

**Results:** Mobility (0.041, 95% CI: 0.033 to 0.048), testing (-0.008, 95% CI: -0.015 to -0.001), and vaccination (quadratic term: 0.004, 95% CI: 0.003 to 0.005, linear term: -0.130, 95% CI: -0.161 to -0.099) were all associated with COVID-19 incidence. For vaccination rate, the decrease of number of cases demonstrated a U-shaped curve, while mobility showed a positive association and testing showed an inverse association with COVID-19 incidence. Mobility restriction was effective during all three periods – March 28, 2020 to November 24, 2021 (0.009, 0.048, and 0.026 respectively). Testing was effective during the second and third period – October 8, 2020 to November 24, 2021 (-0.036, and -0.006 respectively).

**Conclusion:** Mobility restriction and testing were effective even in the presence of vaccination. This shows the positive value of mobility restrictions, testing, and vaccination from the health system perspective on COVID-19 prevention and control, especially with continual emergence of variants in India and globally. At the same time, this health system gain must be balanced with the challenges in the delivery of non-COVID health services and broader socio-economic impact in deciding the prolonged continuance of mobility restriction.

## Introduction

The emergence of coronavirus disease 2019 (COVID-19) has had a colossal impact on public health and the economy worldwide^1^. There has been much effort to control the spread of the disease through measures such as mobility restriction and quarantine^2^. After vaccination rollout began in December 2020, many countries focused on vaccination as its major control measure, while keeping some level of social distancing and quarantine policy^3^. Overall, the pandemic itself, and the interventions implied to control the pandemic both continue to have widespread health, economic and social impact^4-6^.

India has experienced two major outbreaks in September 2020 and May 2021, including a widespread infection of the Delta variant^7,8^. However, it is currently managing the number of daily incidence to under 5 cases per million people (4.91 cases per 1 million people as of December 21, 2021)^7^. Despite relatively low vaccination rate (40% of the population fully vaccinated – two doses – as of December 21, 2021), India has been able to successfully control the number of cases^7^.

With the presence of multiple strategies to control the pandemic, there is a need for a comprehensive assessment of the combined effects of the non-pharmaceutical and pharmaceutical control measures. While there are studies assessing the individual effects of policies or projecting the effect of policies through modelling methods, there are not enough studies that show the combined effect of non-pharmaceutical and pharmaceutical interventions using empirical evidence^9-11^.

Therefore, this study aims to examine the combined effect of interventions in India that have been able to successfully control the spread of the epidemic. Multiple regression analysis was performed to evaluate the combined effect of mobility restriction, testing, and vaccination within the period when vaccination was available. An additional analysis on the entire pandemic period was performed to demonstrate how the effects of non-pharmaceutical interventions (mobility restriction and testing as the proxy of trace-testing-isolation-quarantine policy) changed over time.

## Methods

### Study Population and Data

Data on COVID-19 cases, testing, and vaccination from January 25, 2020 to November 25, 2021 were obtained from publicly available datasets in Our World In Data (published on http://ourworldindata.org on November 26, 2021)^7^. Datasets were linked from official sources and aggregated on the website. Case data on the website were linked from the Johns Hopkins University dashboard. Testing data were linked from official government data obtained from the Indian Council of Medical Research. Vaccination data were linked from the Government of India. Data on population mobility from February 15, 2020 to November 25, 2021 were obtained from Google Community Mobility Reports^12^.

### Variables

The dependent variable for the study was the log-transformed new cases per million-per day during April 28, 2021 to November 24, 2021. The four independent variables were (1) the 7-day moving average of mobility 7 days prior to the index day, (2) the 7-day moving average of Negative/Positive test ratio 7 days prior to the index day, (3) the effective fully vaccinated number per 100 people 14 days prior to the index day, and (4) the log-transformed new cases per million 14 days prior to the index date. 7-day moving average was used to offset daily differences within a week. A 7-day period was applied for mobility and testing to account for the time between the non-pharmaceutical interventions and its effect on incidence. A 14-day period was applied for vaccination to consider the time needed to develop vaccine-derived immunity. The log-transformed new cases per million 14 days prior to the index date was added to adjust for the bias in incidence caused by the pre-existing infectious population in the community.

Negative-Positive ratio was used as a proxy to represent the rigorousness of tracing-testing-isolation and quarantine at any given period of time. The Negative-Positive test ratio was calculated by dividing the negative cases on a specific date by the number of positive cases on the same date. Negative cases were calculated by subtracting the number of new tests on a day by the number of new cases on a day. More negative cases tested per positive case would represent more exhaustive tracing of positive cases to quarantine the contacts of the positive cases unless there was a high number of testing irrelevant to the tracing process.

The mobility change was provided in the Google Community Mobility Reports as the daily difference in mobility compared to the baseline period (between January 3 and February 6, 2020) for 6 different categories of places(workplaces, transit stations, retailer and recreational places, residential areas, groceries and pharmacies, and parks). The average of three of the place categories which were most susceptible to mobility change due to COVID-19 mobility restriction (retailer & recreational, transit station, and workplaces) was used as a proxy for change in mobility^13^.

For vaccination, an operational definition for effective vaccination was constructed to account for the waning effect of vaccination overtime based on empirical research results. We calculated the waning effect of vaccination as a 20 percent decrease after four months^14^. Using this, the effective vaccination rate was calculated by subtracting the waning effect from the number of fully vaccinated population per 100 people (those who have completed their 2nd dose).

### Statistical analysis

To account for missing data points in testing and vaccination, we used the interpolation method to impute missing data. Data for total number of tests and fully vaccinated population were imputed. For the period when the fully vaccinated population was over 1 percent (April 28, 2021 to November 24, 2021), we applied a multiple regression analysis to estimate the effect each of the four independent variables (the 7-day moving average of mobility 7 days prior to the index day, the 7-day moving average of Negative/Positive test ratio 7 days prior to the index day, the effective fully vaccinated number per 100 people 14 days prior to the index day, the log-transformed new cases per million 14 days prior to the index date) had on the dependent variable (log-transformed new cases per million-per day). Graphs were plotted for each independent variable accounting the effect of the other independent variables. An additional multiple regression analysis on the period when the number of cumulative confirmed cases were over 100 (March 28, 2020 to November 24, 2021) was done to analyze the effects of the two non-pharmaceutical interventions (mobility change and Negative/Positive test ratio) prior to the distribution of vaccinations. The log-transformed new cases per million 14 days prior to the index date was also added as a independent variable to the multiple regression analysis to adjust the effect of the pre-existing infectious population in the community.

## Results

We conducted our analysis based on data from April 28, 2021 to November 24, 2021 on daily COVID-19 infections, mobility, tests, and vaccination in India. The trend in 7-day moving average mobility (figure 1-a.) showed a large range of change (-49.5% ∼ +13.64%) with a sharp decrease during the early period (April 28, 2021 to May 23, 2021) and a relatively steady increase until the end of the study period (May 23, 2021 to November 24, 2021). Negative/Positive test ratio (figure 1-b.) was stable during May and June and then increased until July and showed a slow increase until the end of the study period (3.41 ∼ 110.2, range: 106.61). The increase in Negative/Positive test ratio indicates that testing of COVID-19 became more rigorous over the study period. There was an increase in effective vaccination rate from 1.03 to 24.18 during the study period of April 28, 2021 to November 24, 2021 (figure 1-c). The incidence of COVID-19 in the population, represented by the number of new cases per million 14 days prior to the index date, showed an increase in the early period and an overall decrease throughout the study period with small fluctuations (Figure 1-d.).

**Figure 1:**
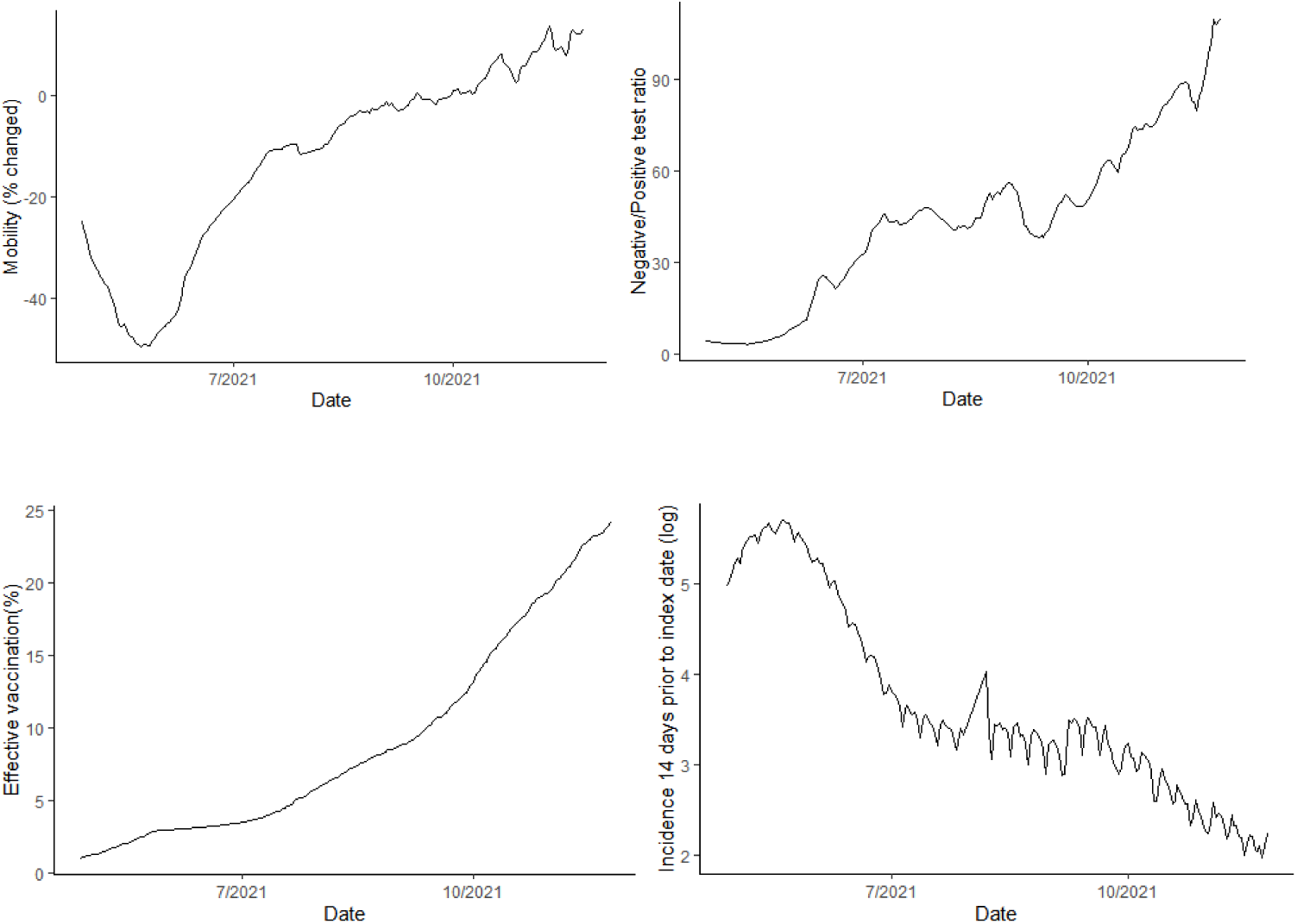
Time trends for mobility, testing, vaccination, and incidence 14 days prior to index date during the COVID-19 pandemic in India during April 28, 2021 to November 24, 2021. Time series graph of mobility (1-a), Negative/Positive test ratio (1-b), effective vaccination (1-c), and incidence 14 days prior to index date (1-d). Negative/Positive test ratio was constructed by dividing the number of negative test results by number of daily new incidents on each day. Effective vaccination was constructed by subtracting the waning vaccination efficacy from the percentage of population that was fully vaccinated (completed 2^nd^ dose of COVID-19 vaccination).

**Figure 2:**
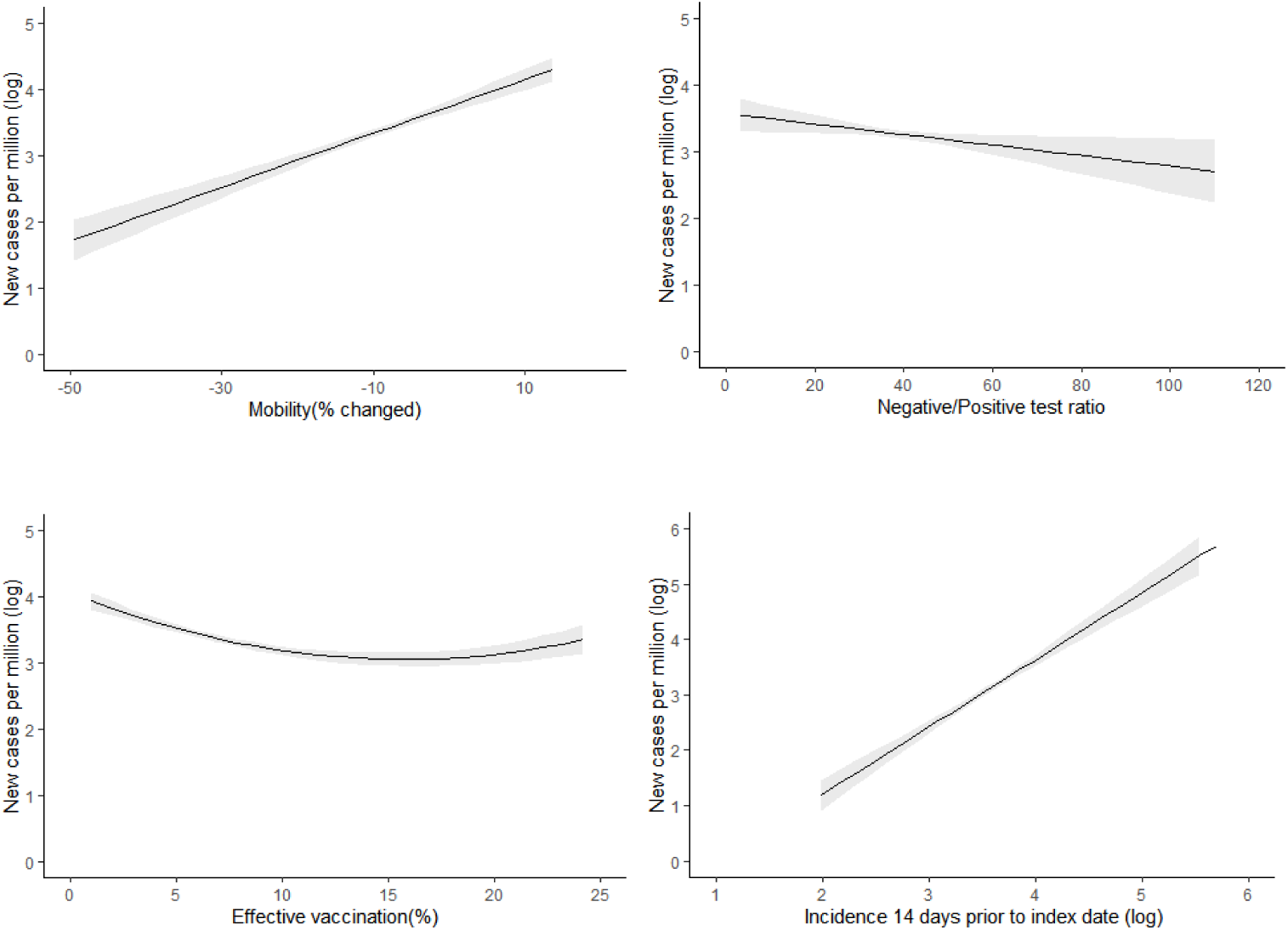
Association of mobility, testing, vaccination, and incidence 14 days prior to index date trends with COVID-19 incidence during the COVID-19 pandemic in India post-vaccination. Multiple regression analysis on post-vaccination period (April 28, 2021 to November 24, 2021) showing the effect of mobility (2-a), Negative/Positive ratio (2-b), effective vaccination (2-c), and incidence 14 days prior to index date (2-d) on the number of new case per day. Incidence 14 days prior to index date was used as a proxy to account for the prevalence of COVID-19 in the community, which affects the pre-existing infectivity of the population.

We conducted a multiple regression analysis of the combined effect of mobility, Negative/Positive test ratio, effective vaccination rate, and pre-existing COVID-19 infectivity of the population on COVID-19 incidence (number of new cases per million). All four independent variables showed statistically significant (p-value<0.05) correlations with the dependent variable. The 7-day moving average mobility from 7 days prior to the index date (0.041, 95% CI: 0.033 to 0.048), and incidence of COVID-19 in the population 14 days prior to the index date (1.216, 95% CI: 1.046 to 1.387) was linearly positively associated with the number of new cases per million on the index date. This indicates that increase of mobility in the previous week is correlated with the increase of number of cases in the following week. The 7-day moving average of Negative/Positive test ratio from 7 days before the index date showed a linearly inverse relationship with new cases per million on the index date (-0.008, 95% CI: -0.015 to -0.001). This shows that rigorous testing in the prior week led to a decrease in cases in the following week. Effective vaccination 14 days prior to the index date showed a U-shaped relationship with the number of new cases per million (quadratic term: 0.004, 95% CI: 0.003 to 0.005, linear term: -0.130, 95% CI: -0.161 to -0.099). This finding indicates that higher effective vaccination rates were associated with fewer cases, even after considering the effect of mobility and Negative/Positive test ratio. Together, these four variables explained 95.6% of the variance of the dependent variable (Multiple R^2^ = 0.9555).

An additional period of March 28, 2020 to November 24, 2021 was analyzed to see the association between mobility change and Negative/Positive test ratio with the number of new cases per million separately during a longer period of time (figure 3). For the analysis, we divided the pre-vaccination pandemic period (defined as the dates with at least 100 cumulative cases of COVID-19) into two periods to see change in effects in the early and late periods of the pandemic. As a result, the entire pandemic period was divided into three periods – 1st period: March 28, 2020 ∼ October 7, 2020, 2nd period: October 8, 2020 ∼ April 27, 2021, 3rd period: April 28, 2021 ∼ November 24, 2021. During March 28, 2020 ∼ October 7, 2020, mobility had a statistically significant positive correlation with the number of new cases per million (0.009, 95% CI: 0.003 to 0.014), while Negative/Positive test ratio did not show a statistically significant correlation with the number of new cases per million (p-value = 0.69497). During October 8, 2020 ∼ April 27, 2021, mobility showed a statistically significant positive correlation with COVID-19 incidence (0.048, 95% CI: 0.032 to 0.064) and Negative/Positive test ratio showed a statistically significant inverse correlation with the dependent variable (-0.036, 95% CI: -0.0452 to - 0.026). During April 28, 2021 ∼ November 24, 2021, which was the period when vaccination was present, this trend continued with mobility showing a positive correlation (0.026, 95% CI: 0.018 to 0.034) and Negative/Positive test ratio showing an inverse correlation on the number of cases per million (-0.006, 95% CI: -0.011 to -0.002). The results from October 8, 2020 ∼ April 27, 2021 were consistent with the analysis for the post-vaccination period while data from March 28, 2020 ∼ October 7, 2020 did not show significant association between Negative/Positive test ratio and the COVID-19 incidence of cases per million.

**Figure 3:**
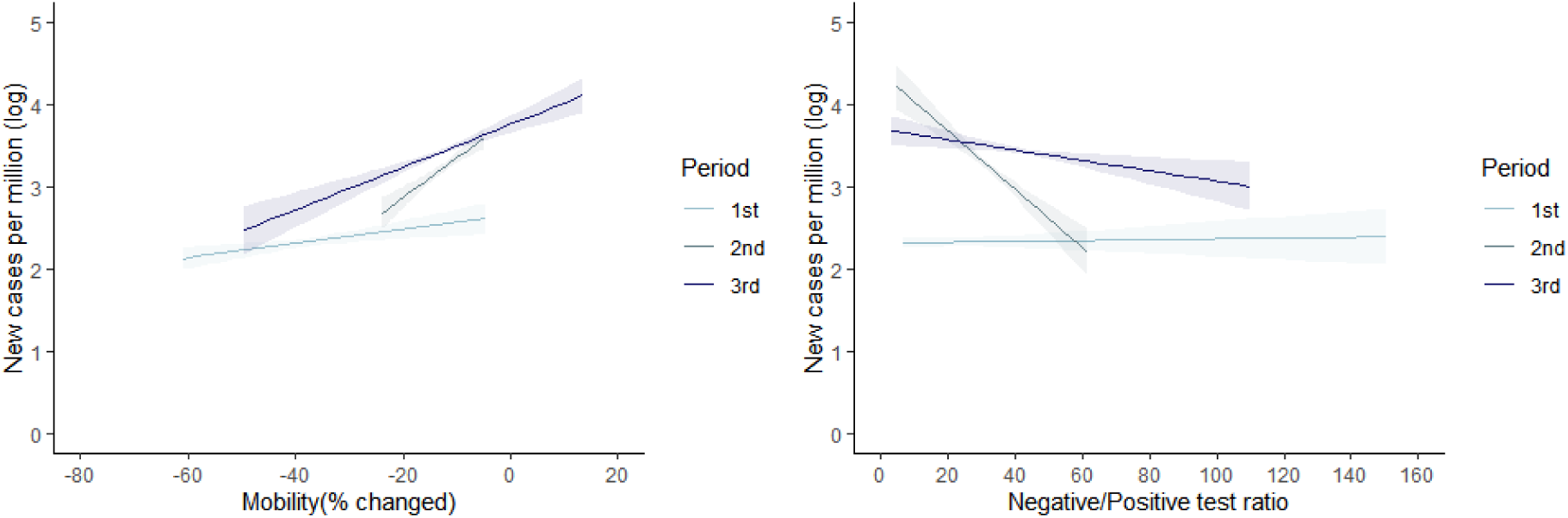
Association of mobility and testing with COVID-19 incidence during the COVID-19 pandemic in India pre- and post-vaccination. Multiple regression analysis on entire period (2020/3/28-2021/11/24) showing the effect of mobility (3-a) and Negative/Positive test ratio (3-b) on the number of new cases per day. (1st period: March 28, 2020 ∼ October 7, 2020, 2nd period: October 8, 2020 ∼ April 27, 2021, 3rd period: April 28, 2021 ∼ November 24, 2021). The effects of mobility and Negative/Positive test ratio differed by period. During the early pre-vaccination period (March 28, 2020 to October 7, 2020), Negative/Positive test ratio did not show a significant correlation with COVID-19 incidence.

## Discussion

Using multiple regression analysis, we have demonstrated that mobility restrictions and testing are effective interventions in prevention and control of COVID-19 even in the presence of vaccination in India. Mobility restriction and rigorous tracing-testing-isolation and quarantine both contributed to a smaller number of daily cases. Changes in the effects of the two major non-pharmaceutical interventions (mobility restriction and testing as a proxy of trace-test-isolation-quarantine) over time were also observed in the study. Mobility restrictions and testing were more effective in the second half of the pre-vaccination period than the first half.

To our knowledge, while there were studies to demonstrate the individual effectiveness of various non-pharmaceutical interventions on the incidence of COVID-19, there was a gap in knowledge on the real-world combined effect of non-pharmaceutical interventions (mobility restriction and testing) and pharmaceutical interventions (vaccination). We conducted a multiple regression analysis on real world data to demonstrate the effectiveness of mobility restriction and testing in the presence of vaccination. In doing so, waning vaccination efficacy, which is an emerging public health concern, was also taken into consideration.

We have limitations in this analysis. Although the analysis was constructed to ensure a temporal relationship between the independent variables and dependent variable by a week, it is hard to conclude a causal relationship yet. We used daily statistics based on testing and were thus unable to analyze the number of quarantined people per case directly, which is an important element of the tracing-testing-isolation and quarantine policy. This may underestimate the impact of tracing-testing-isolation and quarantine when tests are conveniently done decoupled with trace and quarantine. Additionally, we conducted the analysis on a national level. So, the heterogeneity between states in the COVID-19 prevention and control policies and practices is not taken into account in this analysis.

The emergence of new variants, waning vaccination effectivity, and continued breakthrough infections are possible contributing factors to the persistent effectiveness of mobility restrictions and testing. The emergence of highly contagious variant viruses, such as the Delta and Omicron continue to threaten the health system^15,16^. Also, studies show that the effect of currently available vaccinations wane over time, both in serology-based laboratory studies and in empirical studies^17,18^. Studies have also reported breakthrough infections that occur among fully vaccinated population (those who have completed their 2nd dose)^19^. In the presence of such circumstances, high vaccination rates alone do not seem to ensure low number of cases, as seen in the case of South Korea, where over 80% of the population is fully vaccinated, but is nonetheless experiencing record-high numbers of daily new cases^7^.

Thus, the non-pharmaceutical interventions of mobility restrictions and testing maintain their effectiveness in the suppression of the COVID-19 pandemic. This in consistent with research on the early phases of the pandemic that demonstrate that mobility restriction and tracing-testing-isolation and quarantine methods were implemented to suppress the spread of SARS-CoV-2 virus and were highly effective^20^. In Hong Kong, non-pharmaceutical interventions including physical distancing and isolation and quarantine were associated with reduced transmission of COVID-19^21^.

In conclusion, mobility restriction and testing were effective in combination with vaccination on COVID-19 prevention and control in India. While this shows the positive value of mobility restrictions, testing, and vaccination from the health system perspective on COVID-19 prevention and control, especially with the continual emergence of variants of concern in India and globally, this health system gain must be balanced with the challenges in the delivery of non-COVID health services and broader socio-economic impact on the society to decide for or against sustaining the mobility restrictions for a prolonged period of time. With the increasing likelihood of COVID-19 becoming an endemic disease, it is becoming critical to assess the impact of the prevention and control measures on the COVID-19 pandemic within the broader role of the health system in the organization and delivery of all preventative and curative health services while also not impacting the economy and the psychological well-being of the population.

## Data Availability

The datasets analyzed during the current study are available in Our World in Data and Google Mobility Report, https://ourworldindata.org/coronavirus, https://www.google.com/covid19/mobility/.

https://www.google.com/covid19/mobility/

## Abbreviations

COVID-19: Coronavirus disease 2019
CI: Confidence interval

## Declarations

### Ethics approval and consent to participate

Not applicable

### Consent for publication

The corresponding author had full access to all of the data and the final responsibility to submit for publication.

### Availability of data and materials

The datasets analyzed during the current study are available in Our World in Data and Google Mobility Report, https://ourworldindata.org/coronavirus, https://www.google.com/covid19/mobility/.^7,12^ Neither the funders nor the collectors/curators/delivers of the data bear any responsibility for the analyses or interpretations presented here. The statistical code is available from the corresponding author.

### Authors’ contributions

JS and JO conceptualized. JS did the statistical analysis and led the design and writing of the paper. KL contributed to the design, statistical analysis, and interpretation of the analysis. KA advised on the graphical presentation and contributed to the writing of the paper. JO oversaw the design, statistical analysis, interpretation, and writing of the paper. All authors read and approved the final manuscript.

